# When clinical prediction models do not generalize: a simulation study in liver transplantation

**DOI:** 10.64898/2026.03.19.26348780

**Authors:** Damian Brülhart, Giulia Magini, Antonia Schäfer, Simon Schwab, Ulrike Held

**Affiliations:** Department of Biostatistics at the Epidemiology, Biostatistics and Prevention Institute, University of Zurich; Division of Transplantation, Geneva University Hospitals; General Internal Medicine Division, Geneva University Hospitals; Research and Quality Management, Swisstransplant; Center for Reproducible Science and Research Synthesis, University of Zurich

**Author notes:** These authors contributed equally (Simon Schwab, Ulrike Held). **Corresponding author:** Prof. Dr. Ulrike Held, Department of Biostatistics, Epidemiology, Biostatistics and Prevention Institute, University of Zurich, Hirschengraben 84, 8001 Zurich, Switzerland.

## Abstract

**Objectives:** Clinical prediction models estimate the risk of a future outcome in patients. Such models are often externally validated using independent datasets; however, even when a model has been rigorously validated in a new setting and patient population, its performance across other clinical settings remains unclear. Therefore, we systematically evaluated model performance and clinical utility across diverse patient populations to quantify the limits of transportability.

**Methods:** Using liver transplantation as an example, we used the UK donation-after-circulatory-death (DCD) risk score and descriptive statistics from Swiss DCD liver transplant populations to simulate realistic target populations with varying donor and recipient characteristics. The risk score’s ability to predict one-year graft failure was evaluated using calibration intercept, calibration slope, area under the receiver operating characteristic (ROC) curve, and net benefit.

**Results:** The UK DCD Risk Score’s performance depended heavily on the simulated population characteristics. While the score performed adequately in settings similar to those where it was derived, it was not satisfactory in others.

**Discussion:** The study showed, using a risk score in liver transplantation as an example, that the application of a prediction model can be limited in certain external populations when they differ, and that its transportability in new settings is not guaranteed.

**Conclusion:** This study highlights the importance of external validation of clinical prediction models to determine transportability to various target populations. Their application requires careful consideration and potential model re-estimation.

## INTRODUCTION

### Background

Liver transplantation is the therapy of choice for patients with end-stage liver diseases. The transplantation improves survival and quality of life of patients while also having a beneficial impact on public health and the socio-economic burden of organ failure [1]. Donation after circulatory death (DCD) describes organ transplantations after the donor’s death has been confirmed according to circulatory criteria prior to brain death. Due to the loss of circulation, these organs generally are more susceptible to ischemic-reperfusion injuries than those donated after brain death (donation after brain death; DBD) or from a living donor. The large supply-demand gap has led to many countries allowing the opportunity for DCD [2]. However, due to the fragility of these organs, proper methods for selection and allocation must be in place to ensure the safety of the recipient and the longevity of the graft.

Clinical prediction models are a cornerstone of personalized medicine and can support clinical-decision making by estimating a patient’s risk of a specific outcome. Many prediction models have been developed to guide the decision-making processes in transplant medicine, among them the UK DCD Risk Score [3]. The UK DCD Risk Score is a prognostic clinical prediction model used to estimate the risk of one-year graft failure for a possible combination of donor and recipient. The model is based on seven donor-and-recipient key variables to determine a risk score and to classify the combination into a low-risk, high-risk or futile group. The model was trained and validated on adult DCD cases from the United Kingdom (UK) [3].

The Swiss target population includes the donors and recipients in liver transplantations in Switzerland. The population used for training and validation of the UK DCD Risk Score is suspected to be different from the target population in Switzerland due to regulatory differences and differences in target population characteristics. For example, in Switzerland almost no retransplantations (recipient receiving a second liver transplant after the first has failed) are conducted, which is a very important factor in the UK DCD Risk Score. Furthermore, there have been discussions about potential statistical flaws in the UK DCD Risk Score [4] and its usefulness in certain population settings [5].

### Objectives

The performance of clinical prediction models across varying patient populations remains unclear. This is particularly relevant in solid organ transplantation, where patients’ characteristics, clinical processes, and follow-up care may differ across hospital transplant centres and countries.

The aim of the study was to validate the UK DCD Risk Score in different simulated population settings and thereby quantify the generalizability and transportability of the clinical prediction model. The varying target populations were based on real-world data from Switzerland.

## METHODS

### The UK DCD Risk Score

The UK DCD Risk Score aims to predict graft-failure in the first year after liver transplantation. It is based on a logistic regression model and transformed into a point system between 0 and 27 with lower values indicating lower risk of one-year graft failure. The model was developed using data from DCD liver transplantations in the UK. The data included 1153 patients from a national UK cohort for development and data from two different cohorts from the US and the UK with 1861 and 315 DCD transplantations for validation, respectively [3].

The UK DCD Risk Score is partly used in Switzerland to assess the risk of liver graft failure for certain donor-recipient combinations in certain situations during the organ allocation process. The seven predictors for the UK DCD Risk Score are donor age (D.age), donor BMI (D.BMI), functional donor warm ischemia time (FWIT), cold ischemia time (CIT), recipient age (R.age), recipient model for end-stage liver disease (MELD) score (R.MELD) and retransplantation (retp). The continuous variables are di- or trichotomized and points are distributed according to Table 1. An example is given in the Supplemental material.

**Table 1:** The points system for the UK donation-after-circulatory-death (DCD) Risk Score, which ranges from 0 to 27. For each of the factors present below, the corresponding points are rewarded when the condition is met (or zero otherwise). Higher values indicate higher risk of graft failure[3]. D.age: donor age; D.BMI: donor BMI; FWIT: functional donor warm ischaemia time; CIT: cold ischaemia time; R.age: recipient age; R.MELD: recipient lab model for end-stage liver disease (MELD) score; retp: retransplantation

| Factor | Points |
| --- | --- |
| D.age > 60 y | 2 |
| D.BMI > 25 | 3 |
| FWIT $\in (20, 30]$ min | 3 |
| FWIT > 30 min | 6 |
| CIT > 6 h | 2 |
| R.age > 60 y | 3 |
| R.MELD > 25 | 2 |
| retp TRUE | 9 |

The score can be categorized into three groups [3]:

- Score ≤ 5 points: low-risk group, one-year graft survival ≥ 95%
- Score ∈ (5, 10] points: high-risk group, one-year graft survival > 85%
- Score > 10 points: futile group, one-year graft survival < 40%

### Data-generating mechanism

The simulation of the predictor variables was based on descriptive statistics from Swisstransplant. The outcome was always simulated twice – once according to the UK DCD assumptions and once according to the Swisstransplant data. The mechanism is described more closely in the Supplemental material.

### Performance measures

The performance measures used to evaluate the UK DCD Risk Score in the different simulation settings were calibration intercept, calibration slope, net benefit to transplant none, net benefit to transplant all, and the area under the receiver operating characteristic (ROC) curve (AUC).

The calibration intercept and slope were calculated by fitting a linear model with the observed probabilities as response and the predicted probabilities as explanatory variable. A calibration intercept of 0 and a calibration slope of 1 would indicate perfect calibration.

In order to compare the strategy of using the risk score to the strategies of transplanting all, we calculated the differences in net benefit between these strategies with a threshold probability of 80% and the decision rule of only transplanting donor-recipient combinations classified as non-futile [6].

The calculation of the calibration intercept and slope as well as the net benefit are described in the Supplemental material in more detail.

The AUC was calculated using the pROC package in R [7,8]. Perfect discrimination is indicated by an AUC of 1, while 0.5 indicates that the discrimination is no better than chance.

### Simulation design

Each simulated condition was repeated 1000 times. If 1000 iterations did not yield a Monte Carlo standard error (MCse) of less than 0.01 for the AUC, the simulation size would have been increased. We set up four different simulations in which we varied different population characteristics:

- Mean D.age and R.age (fully factorial)
- Mean FWIT and probability for CIT > 6 h (fully factorial)
- Retransplantation probability
- Sample size

In each simulation aspect (each bullet point), the characteristics described were varied, while all other characteristics were kept at baseline. The baseline was the simulation setup mirroring the Swiss target population with sampling parameters according to Supplemental Table 2 and a sample size of 1200, reflecting the sample size used in the development of the UK DCD Risk Score. All simulations were run twice, once with outcome simulation according to the UK DCD dichotomized model and once according to the Swisstransplant model.

### Flowchart

For better understanding of the simulation study, a flowchart was created. The flowchart is shown in Figure 1.

**Figure 1:**
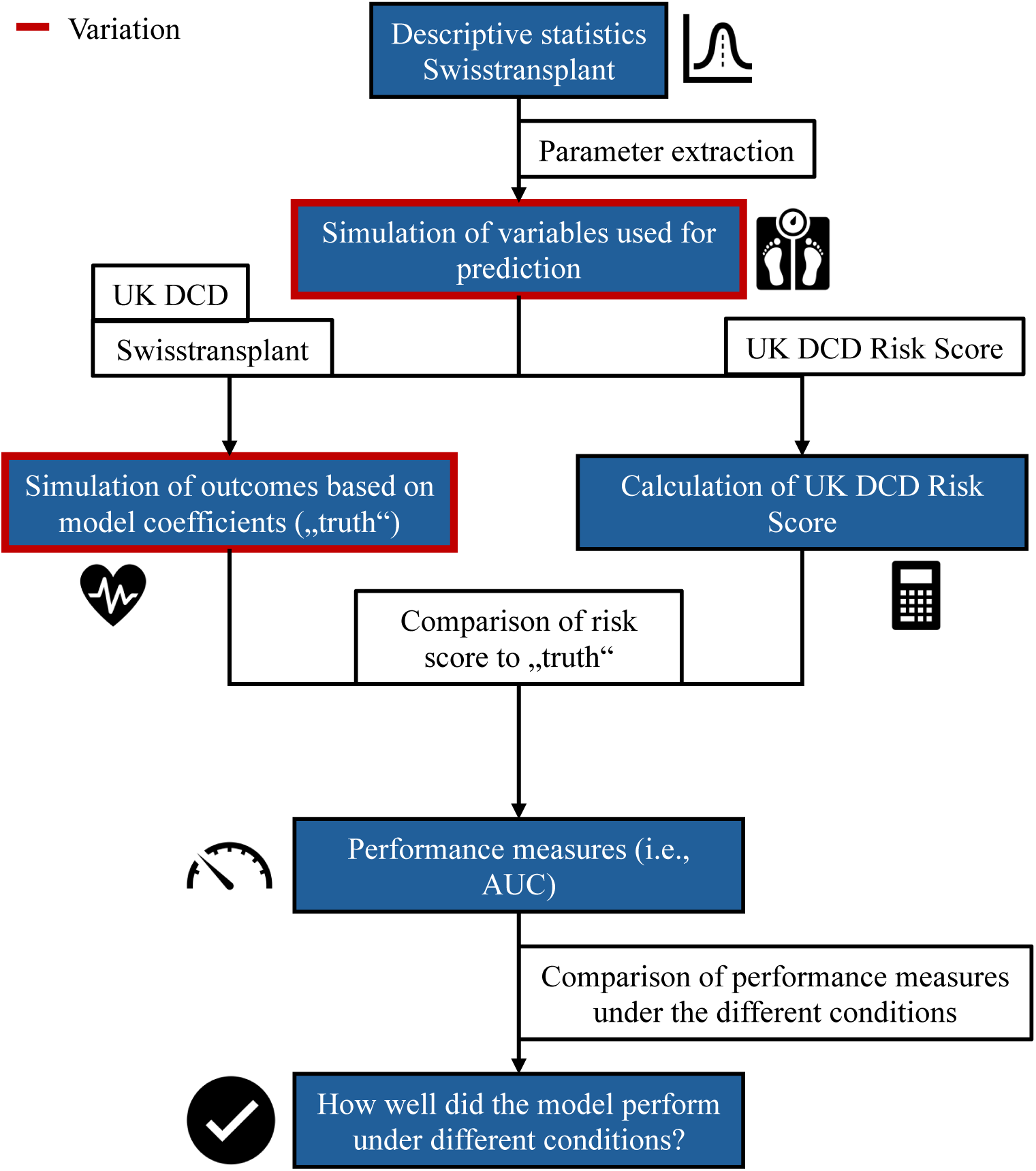
Flowchart of the simulation study. The steps marked by a red frame are the steps where different simulation conditions were used (e.g., different donor or recipient ages, different outcome models, …). DCD: donation-after-circulatory-death; AUC: area under the receiver-operating-characteristic (ROC) curve

### Missingness and non-convergence

Missing values and non-convergence were reported and handled according to current suggestions [9]. The detailed approach is described in the Supplemental material.

### Pre-registration

This simulation study was pre-registered according to the ADEMP (aims, data-generating mechanisms, estimands, methods, and performance measures) pre-registration guideline based on the publication by Siepe et al. (2024) [10]. All versions of the pre-registration document can be found at github.com/dbruel/PreReg_MT.

### Code and data sharing

To ensure reproducibility, all code used to generate the results of the simulation study are publicly available at http://github.com/dbruel/TransPM.

### Ethical approval

Ethical approval was requested from the Ethics Committee of the Faculty of Medicine at the University of Zurich to use the descriptive statistics, correlations and the model based on the Swisstransplant data. The application was reviewed and the committee approved the study (MeF-Ethik-2025-15). Because the study is not a clinical trial, it was not further registered.

### Reporting

This simulation study is reported using the TRIPOD+AI guideline for the development or evaluation of clinical prediction models whenever applicable [11].

### Software

The simulation and all analyses were conducted in R [7] version 4.5.0. The simulation was set up using the SimDesign package [12]. The AUC was calculated using the pROC package [8]. Plotting of the results was achieved using the ggplot2 package [13].

## RESULTS

### Variation of donor and recipient age

The performance of the UK DCD Risk Score under varying mean donor and recipient age in the target population is shown in Figure 2. When the outcome was simulated according to the UK DCD logistic regression model, the UK DCD Risk Score performed best when the mean donor and recipient age in the target population were around 60. For this target population, the prediction model showed the best calibration, the best net benefit and quite high discrimination as reflected in the AUC.

**Figure 2:**
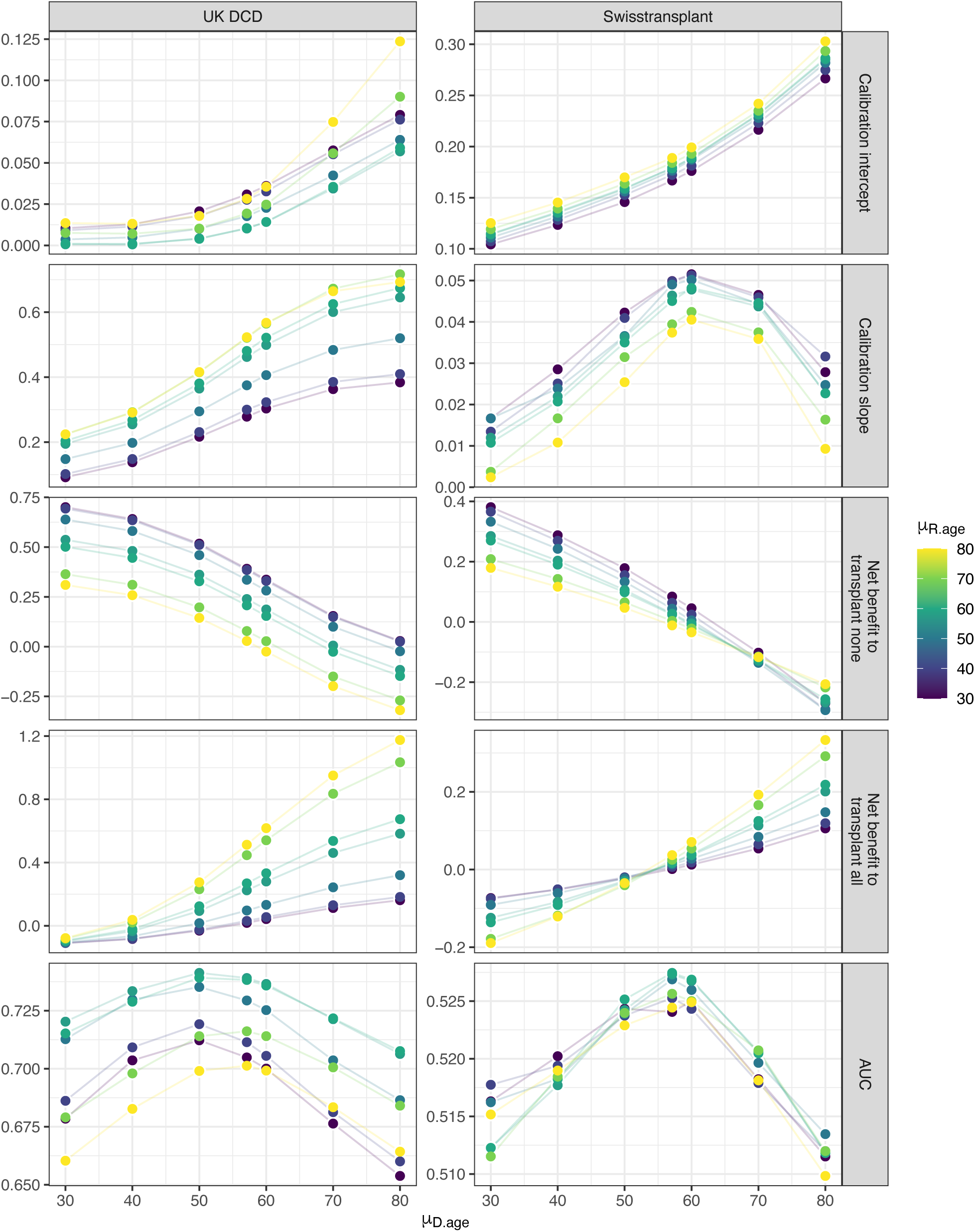
Performance of the UK donation-after-circulatory-death (DCD) Risk Score under variation of donor and recipient age in the target population. The points represent the medians over the iterations. The errorbars were omitted for better visibility. The simulation size for each simulation condition was n_sim_ = 1000. AUC: area under the receiver-operating-characteristic (ROC) curve; D.age: donor age; R.age: recipient age

The UK DCD Risk Score did not perform particularly well in any target population with outcomes simulated according to the Swisstransplant model. However, in a target population with recipient and donor mean ages around 60, the strategy of using the model was not worse than transplanting all or transplanting none which was the case in older and younger target populations. Therefore, also when the outcome was simulated according to the Swisstransplant model, the UK DCD Risk Score performed best in a target population with mean ages around 60 but transplanting all or transplanting none were not inferior strategies.

### Variation of FWIT and CIT

The performance of the UK DCD Risk Score under varying functional donor warm ischaemia time (FWIT) and cold ischaemia time (CIT) with outcome simulation according to the UK DCD logistic regression model or the Swisstransplant model is shown in Figure 3. Under outcome simulation according to the UK DCD model, the UK DCD Risk Score showed the best calibration and discrimination when μ_FWIT_ and P(CIT > 6 h) were low. The net benefit analysis showed that the use of the model was always the superior strategy to transplant all and transplant none.

**Figure 3:**
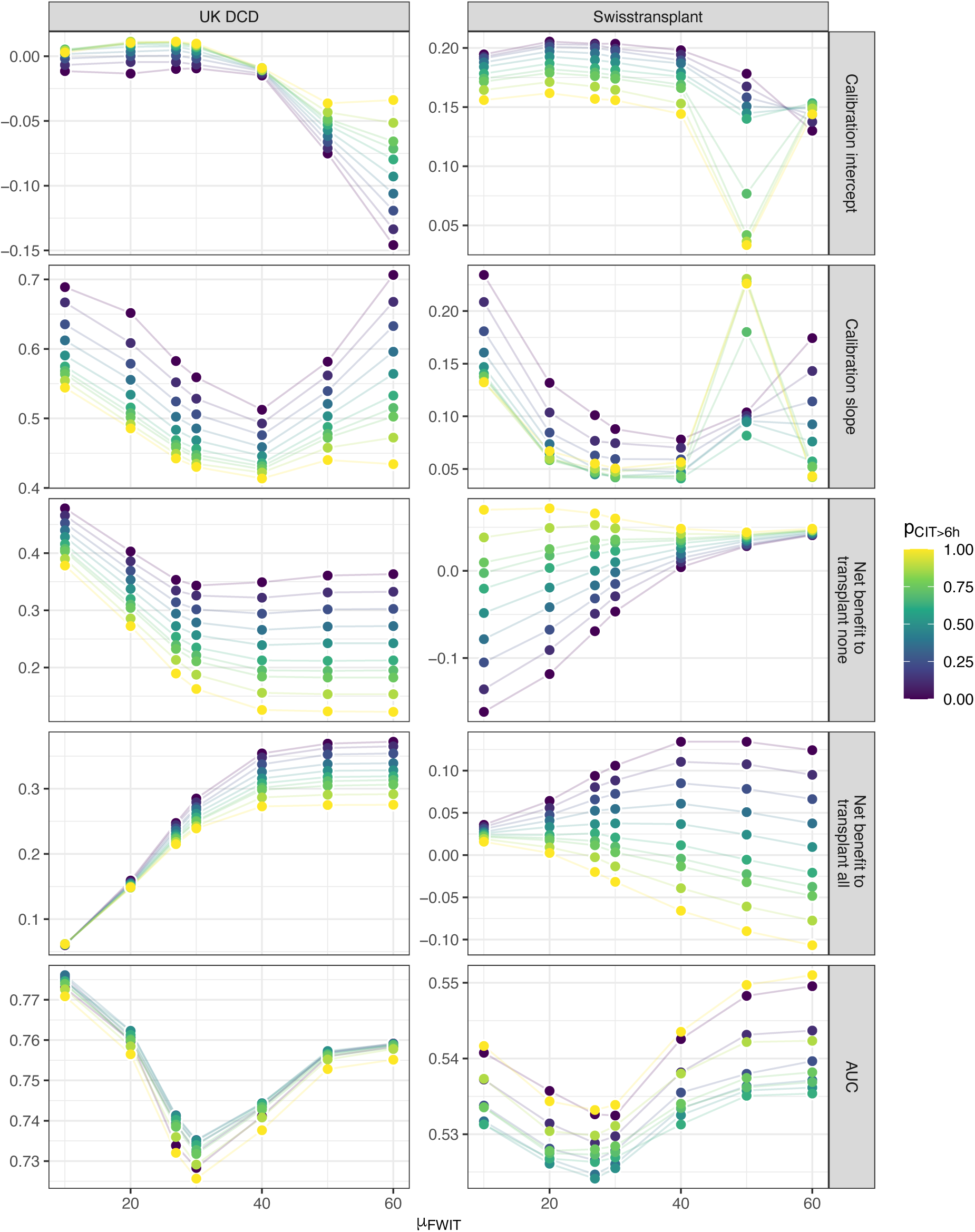
Performance of the UK donation-after-circulatory-death (DCD) Risk Score under variation of functional donor warm ischaemia time (FWIT) and the probability for cold ischaemia time (CIT) greater than 6 hours in the target population. The points represent the medians over the iterations. The errorbars were omitted for better visibility. The simulation size for each simulation condition was n_sim_ = 1000. AUC: area under the receiver-operating-characteristic (ROC) curve; FWIT: functional donor warm ischaemia time; CIT: cold ischaemia time

With outcome simulation according to the Swisstransplant model, the performance of the UK DCD Risk Score was generally bad. The calibration of the UK DCD Risk Score was best with high CIT and μ_FWIT_ = 50 min. However, in these simulated target populations, the transplant all strategy was preferable according to the net benefit analysis. The UK DCD Risk Score was generally slightly better than the other strategies (transplant none and transplant all) when μ_FWIT_ was high and P(CIT > 6 h) low in terms of net benefit. The discrimination of the prediction model in target populations with high μ_FWIT_ was also the highest.

### Variation of retransplantation probability

The performance of the UK DCD Risk Score under varying proportions of retransplantation in the target population is shown in Figure 4. In a population with outcomes simulated according to the UK DCD model, the UK DCD Risk Score showed the best calibration in target populations with approximately 10-20% retransplantations where the calibration slope was high and the calibration intercept close to zero. Discrimination in these populations was also good, reflected by a rather large AUC. From a net benefit perspective, the model outperforms the transplant all and transplant none strategies in all populations (with 0-50% retransplantations). In populations with small retransplantation probabilities, the UK DCD Risk Score did not perform as well.

**Figure 4:**
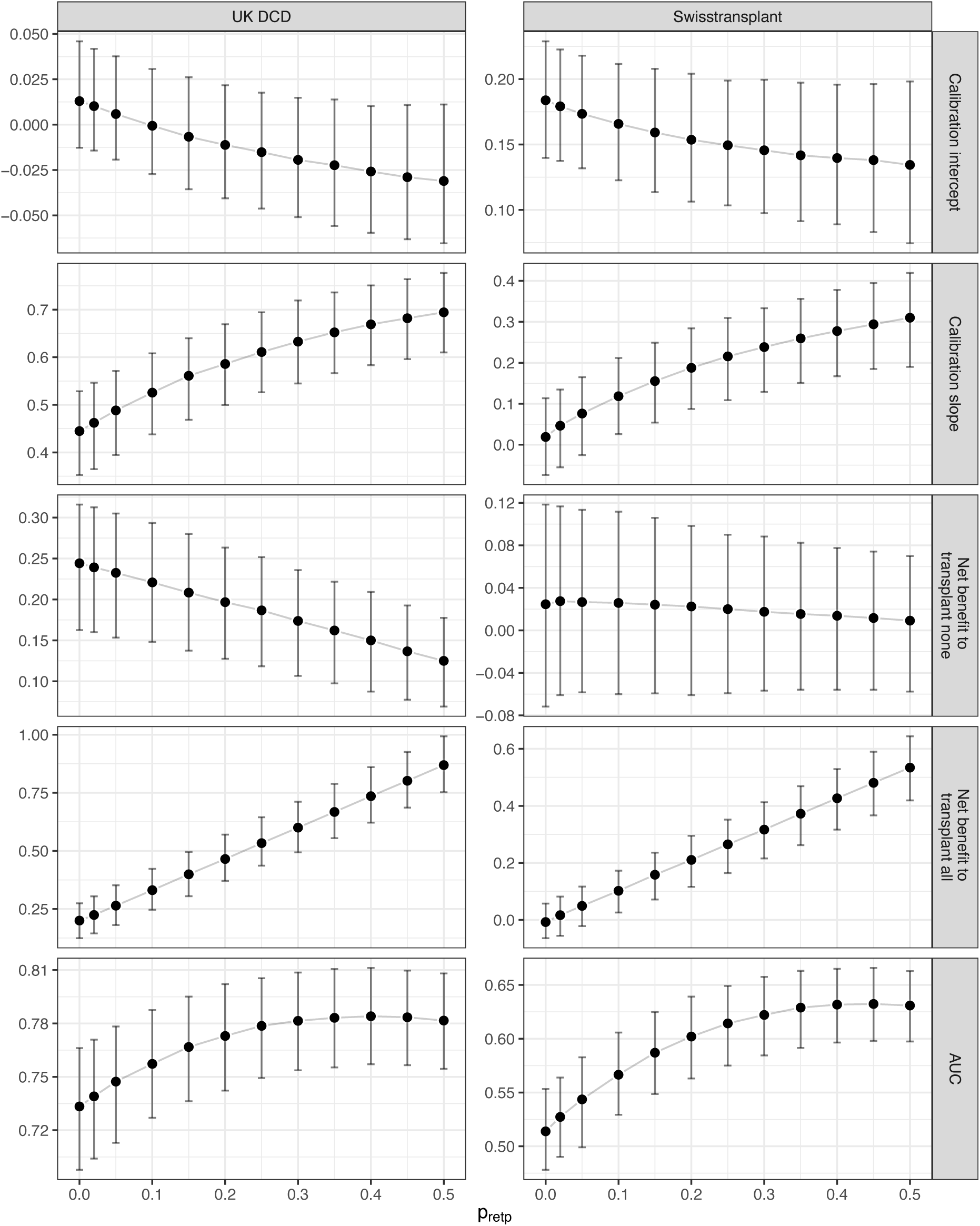
Performance of the UK donation-after-circulatory-death (DCD) Risk Score under variation of probability for retransplantation in the target population. The points represent the medians over the iterations and the error bars the 2.5% and 97.5% quantiles. The simulation size for each simulation condition was n_sim_ = 1000. AUC: area under the receiver-operating-characteristic (ROC) curve; retp: retransplantation

Under outcome simulation according to the Swisstransplant model, the UK DCD Risk Score showed the best calibration and discrimination when retransplantation was more common in the population (30-40%). In these populations, the calibration intercept was closer to zero, the calibration slope closer to one and the AUC much larger than in the populations with few retransplantations. It is important to note that from approximately 20% retransplantations upward, the AUC was above 0.6. According to the net benefit analysis, the use of the model was generally not much better than transplanting none. However, the more retransplantations in the population, the better the model performed compared to transplanting all.

### Validation of the simulated data and variation in sample size

The simulation process did not yield any missing values or non-convergence. Furthermore, the simulated data did not show any major inconsistencies. The validation results are more closely described in the Supplemental material.

The precision was largely affected by sample size, corresponding results can be found in the Supplemental material.

## DISCUSSION

The results of this simulation study show that the performance of clinical prediction models or risk scores (such as the UK DCD Risk Score) are vastly dependent on population characteristics. Previous research has already raised question marks about whether the UK DCD Risk Score is still applicable in external populations [4,5,14] and other models might be good alternatives [15,16]. Additionally, the transportability and need for external validation of prediction models in general has been highlighted before [17–22]. Our findings directly relate to the existing literature and show that the transportability of clinical prediction models depends on external population characteristics. Furthermore, the results highlight the importance of continuous external validation and model updating for adaptation to changing or new target populations and changing transplant protocols (e.g. integration of advanced preservation techniques [23].

The study has some limitations, which should be considered when interpreting the results:

- The data used as a basis for the simulation study only included transplantations which were conducted in Switzerland. This could introduce a bias because the UK DCD Risk Score is sometimes used in the decision-making process. This bias is reduced by the fact that transplantations classified as futile according to the UK DCD Risk Score were still conducted (20-30% depending on the centre).
- The model used for outcome simulation based on the Swiss target population was overfitted and then shrunk by a factor of 0.9. While the shrinkage by a fixed factor generally improves the model used for outcome simulation it can still distort the performance in individual simulated data sets [24,25], leading to potential underfitting.

However, the study also has several strengths:

- The design of the simulation study allowed us to evaluate the UK DCD Risk Score under various population settings. In contrast, classical validation studies utilizing non-simulated data often face limitations in achieving such comprehensive evaluations.
- Basing the simulation study on real-world data lead to more realistic and reliable results compared to traditional simulation studies based solely on theoretical assumptions.
- The validation of the simulated data ensured that the reflection of the intended statistical properties and enhanced the quality of the simulation study.
- Using guidelines such as the pre-registration of the simulation study according to the ADEMP template [10], reporting missingness and non-convergence [9], reporting according to the TRIPOD+AI statement [11] and publishing our code on GitHub ensures the highest standards regarding reproducibility.

This simulation study highlights the importance of external validation and potential model refitting to apply to the new target population. Additionally, we recommend that the performance of a clinical prediction model, which was at some point declared fit-for-use, is continuously monitored and updated as necessary. By adhering to these measures, it can be assured that clinical prediction models always remain robust and reliable in their predictive capabilities. This continuous process of validation and updating ensures that the model adapts effectively to the characteristics of the new population. As a result, it provides accurate, relevant insights that support clinical decision-making. Ultimately, this dynamic approach helps both patients and doctors by offering tailored predictions that enhance diagnostic accuracy, optimize treatment strategies, and improve patient outcomes.

## CONCLUSION

This study highlights the importance of external validation of clinical prediction models to determine transportability to various target populations. Their application requires careful consideration and potential model re-estimation.

## CONTRIBUTORS

DB, SS and UH conceptualized the study. DB wrote the statistical software and performed the statistical analysis. SS and UH validated the analysis. GM, AS and SS provided resources in form of descriptive statistics as foundation for the simulation study. GM and AS provided clinical context. SS and UH were supervisors. DB wrote the initial draft. All other authors participated in writing and reviewing the manuscript. All authors approved the final version of the manuscript.

## COMPETING INTERESTS

The authors declare no competing interests.

## FUNDING

This study received no funding.

## Notes

### Competing Interest Statement

The authors have declared no competing interest.

### Funding Statement

This study did not receive any funding.

### Author Declarations

Ethical approval was requested from the Ethics Committee of the Faculty of Medicine at the University of Zurich to use the descriptive statistics, correlations and the model based on the Swisstransplant data. The application was reviewed and the committee approved the study (MeF-Ethik-2025-15).

