## Supplemental material for "When clinical prediction models do not generalize: a simulation study in liver transplantation"

**Supplemental tables**

*Table 1: Odds ratios (OR) from the UK donation-after-circulatory-death (DCD) model [1] and log odds ratios (logOR) from the Swisstransplant model used for outcome simulation. To counteract potential overfitting in the Swisstransplant model, the coefficients in the table were shrunk by a factor of 0.9.*
*D.age: donor age; D.BMI: donor body-mass-index; FWIT: functional donor warm ischaemia time; CIT: cold ischaemia time; R.age: recipient age; R.MELD: recipient model of end-stage liver disease score; retp: retransplantation*

| **UK DCD** | | **Swisstransplant** | |
|---|---|---|---|
| *Parameter* | *OR* | *Parameter* | *logOR* |
| D.age > 60 y | 11.90 | D.age | 0.026 |
| D.BMI > 25 | 1.93 | D.BMI | -0.003 |
| FWIT > 20 min | 2.93 | FWIT | -0.003 |
| CIT > 6 h | 1.26 | CIT > 6 h | -0.346 |
| R.age > 60 y | 4.15 | R.age | 0.004 |
| R.MELD > 25 | 9.17 | R.MELD | 0.008 |
| retp TRUE | 8.57 | retp TRUE | 1.257 |

*Table 2: Baseline parameters for the simulation as extracted from the descriptive statistics from the Swiss target population.*
*D.age: donor age; D.BMI: donor body-mass-index; FWIT: functional donor warm ischaemia time; CIT: cold ischaemia time; R.age: recipient age; R.MELD: recipient model of end-stage liver disease score; retp: retransplantation*

| **Variable** | **Parameters** |
|---|---|
| D.age | $\mu = 57.1$, $\sigma = 16.1$, $\rho(D.age, D.BMI) = 0.1$ |
| D.BMI | $\mu = 26$, $\sigma = 4.3$, $\rho(D.age, D.BMI) = 0.1$ |
| FWIT | $\mu = 26.9$, $\sigma = 9.4$ |
| CIT | P(CIT > 6 h) = 0.702 |
| R.age | $\mu = 57.8$, $\sigma = 9.9$ |
| R.MELD | $\lambda = 14.7$ |
| retp | P(retp) = 0.02 |

**Example for the calculation of the UK DCD Risk Score**

To calculate the UK DCD Risk Score for a certain donor-recipient combination, assume the recipient is 65 years old and already received a transplant previously (retransplantation TRUE). Otherwise, there are no risk factors present. The score would then be calculated as 3 + 9 = 12 points, which would classify the donor-recipient combination into the futile group with a one-year graft survival of less than 40%.

### Data-generating mechanism

The distributions used for sampling were determined based on descriptive statistics from DCD transplantations in Switzerland between 2013 and 2023. D.age and D.BMI were expected to be correlated and therefore simulated from a truncated multivariate normal distribution bound in $[0, \infty)$ to exclude negative values. The correlation was determined based on the descriptive statistics as 0.1. FWIT and R.age were sampled from truncated normal distributions bound in $[0, \infty)$. CIT and retp were sampled from Bernoulli distributions because CIT was only available dichotomized with a cut-off of 6 hours. R.MELD was sampled from a truncated Poisson distribution bound in [6, 40].

The outcome (one-year graft failure) was simulated using a Bernoulli distribution with the probability for one-year graft failure estimated for each simulated participant. This probability was estimated using two different models: the UK DCD Risk Score logistic regression model [1] and our own Swisstransplant model. The Swisstransplant model based on data from the Swiss target population did not include a sufficient amount of events (54 events) for the amount of estimated coefficients. Therefore, we applied a shrinkage factor of 0.9 to its coefficients [2]. The coefficients of both models are reported in Supplementary Table 1. The estimated probability for one-year graft failure was calculated as

$$\widehat{p_1} = \frac{1}{1+\exp(-\beta X)},$$

where β are the log odds ratios and $X$ are the predictor values for a certain participant.

The intercepts ($\beta_0$) were chosen such that the overall prevalence of one-year graft failure was about 20% when mirroring the conditions of the Swiss target population because this was the percentage in the Swiss target population.

The generated data was validated with respect to its population characteristics. This included checking the distributions of the simulated data and making sure that sampling from the truncated distributions did not largely affect its location and scale measures (e.g., a sample from a normal distribution with $\mu = 0$ bound in $[0, \infty)$ will have mean > 0). Additionally, we verified that whenever the simulated data mirrors the Swiss target population characteristics, the chosen intercept leads to a one-year graft failure prevalence of 20% in the population.

### Handling missingness and non-convergence

All simulation repetitions were monitored for missingness, failures, and non-convergence. Missingness was defined as any simulation run that did not produce a valid output due to algorithmic failure or non-convergence. The frequency and proportions of such occurrences were systematically recorded for each method and simulation condition. If missingness were to occur, we would have evaluated its potential impact on the results and considered whether it was related to specific performance measures or simulation scenarios. Our approach to handling missingness was aligned with the primary goals of the study: if missingness was minimal and randomly distributed, affected runs were excluded from summary statistics; if missingness was substantial or systematically related to certain conditions, we conducted sensitivity analyses to assess its influence on the findings. All results of the simulation study were interpreted in light of the distribution of missing values and non-convergence over the simulation conditions [3].

**Calculation of performance measures**

*Mapping of the UK DCD Risk Score on the probability scale for the calibration curve*: The predicted UK DCD Risk Score was mapped onto the probability scale by dividing the midpoints of the respective low-risk, high-risk and futile group by 27 (e.g., if a simulated participant belonged to the low-risk group, the predicted probability would be 2.5/27 ≈ 0.09). The observed probabilities were calculated as the prevalence of simulated outcomes in each group.

*Net benefit*: The net benefit can be calculated as

$$NB = \frac{TP}{N} - \frac{FP}{N} \cdot \frac{p_t}{1-p_t} \text{ [4]},$$

where *TP* is the number of true positives (participants selected for transplantation without one-year graft failure), *FP* the number of false positives (participants selected for transplantation with one-year graft failure), *N* the total sample size and $p_t$ the threshold probability. The donor-recipient combinations selected for transplantation were defined as the combinations which were classified as non-futile according to the UK DCD Risk Score. The threshold probability is used as a conversion rate of how the harms relate to the benefits of the strategy (selection using model, transplant none or transplant all) and can be defined as the minimum success probability a surgeon would want to still conduct the transplantation. After careful consultation with transplantation experts at Swisstransplant, we defined this probability as 80%. We calculated the difference in net benefit between the model and a transplant all and a transplant none strategy (where the latter is just the net benefit of using the model). A good model would have a high net benefit compared to both the transplant none and transplant all strategies. A model worse than the other strategies would have a negative net benefit compared to the transplant none and transplant all strategies.

**Validation of the simulated data**

The simulation process did not yield any missing values or non-convergence. Significant deviations from the desired location and scale measures were only observed in D.age and FWIT. When D.age was simulated with $\mu = 30$, the mean age was slightly above 30 and the standard deviation lower than expected. When FWIT was simulated with $\mu = 10$ or 20, the mean was also slightly larger and the standard deviation slightly lower than expected. These deviations were associated with the vicinity to the boundaries of the truncated distributions used for sampling at 0 and were taken into account for the interpretation of the results. For all other variables used for prediction, the distributions did not deviate from the targets.

### Variation of sample size results

The performance of the UK DCD Risk Score under varying sample sizes between 10 and 2000 showed the effect of having more or less participants in the validation study. The results of the simulation are shown in Figure 1. The performance measures were independent of the sample size. However, the smaller the sample size, the larger was the chance to receive extreme results by chance, which was reflected in wide ranges between the 2.5 and 97.5% quantiles of the simulation results.

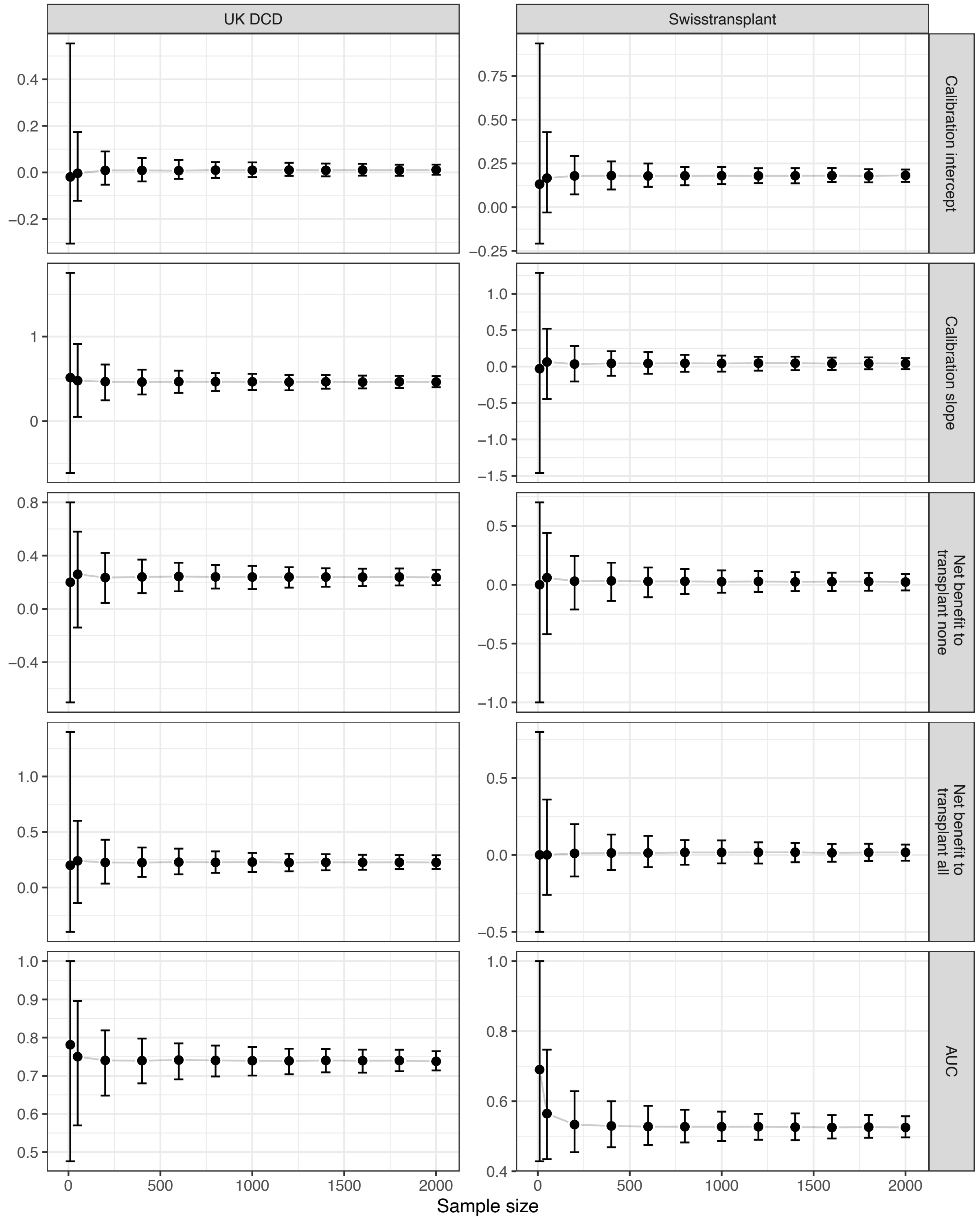

*Figure 1: Performance of the UK donation-after-circulatory-death (DCD) Risk Score under variation of sample size. The points represent the medians over the iterations and the error bars the 2.5% and 97.5% quantiles. The simulation size for each simulation condition was nsim = 1000.*
*AUC: area under the receiver-operating-characteristic (ROC) curve*
